# A health world model for quantitative reasoning on longitudinal health profiles

**DOI:** 10.64898/2026.10.03.26364142

**Authors:** Hongcheng Guo, Lingran Song, Peng Ren, Jinghong Han, Yuxin Wang, Haoran Li, Jiebi Deng, Jiali Wang, Jianfeng Feng, Weikang Gong, Wei Cheng

## Abstract

Medical large language models have shown promise in medical knowledge retrieval and clinical reasoning but remain limited in quantitative reasoning over longitudinal health data, partly due to the lack of effective data construction and model training strategies tailored to large-scale prospective cohorts. Here, we present HealthFound, a health world model trained on 12.4 million constructed examples, including 14-year longitudinal biomedical records from 501,936 participants and datasets for medical knowledge and general reasoning. We propose a three-stage training framework comprising progressive curriculum supervised fine-tuning, rejection-sampled retention fine-tuning, and task-verifiable policy optimization. This enables the model to effectively parse heterogeneous quantitative data and perform robust quantitative reasoning about incident diseases and health trajectories. HealthFound achieves the highest mean accuracy across eight public medical benchmarks. On the task of predicting 1,074 future disease onsets, it outperforms the closest large language model by approximately 10 percentage points on average. In two independent external validations (MIMIC-IV and NHANES), it demonstrates superior zero-shot generalization, achieving improvements of over 5.2% across 417 disease endpoints. The model also achieves top results on tasks involving literature-derived question answering and quantitative indicator imputation. Furthermore, HealthFound generates interpretable reasoning rationales that are consistent with established biomedical knowledge. Collectively, our results demonstrate that HealthFound enables medical LLMs to evolve from static knowledge repositories into dynamic quantitative reasoning engines over longitudinal health data.

## Introduction

Medical large language models (LLMs) have made promising progress on medical examinations, clinical question answering and other forms of clinical reasoning^1,2^. This progress is seen in both general-purpose LLMs and models adapted to medical specialties^3,4^. Much of their competence stems from pretraining or fine-tuning on biomedical literature^5^, clinical notes^6^ and medical instruction data^7,8^. These data are well suited for learning medical facts, interpreting clinical vignettes^9^ and producing plausible explanations^3,4,10^.

However, several critical gaps remain. A model may know how diabetes or heart failure is described, but this knowledge does not establish whether it can interpret an individual’s complex routine measurements—such as vital signs, blood tests or imaging results—and reliably infer later disease outcomes^11^. It also remains unclear how reliably such models can infer an unobserved measurement from the rest of a health profile. Beyond individual disease predictions, another challenge is whether these models can anticipate how a person’s health may evolve, including which clinical events might occur and when.

These limitations reflect the nature of the training signal. Biomedical literature, clinical notes and examination questions are predominantly qualitative and descriptive. They encode concepts and clinical decisions, but provide relatively little supervision over how measurements covary across individuals, how they change over time, or how pre-disease profiles relate to later outcomes. We posit that repeated quantitative observations linked to subsequent outcomes^12^ could supply this missing signal, allowing a language model to learn the statistical structure underlying evolving human health states^13,14^. This provides a foundation for a health world model that learns relationships between past observations, current health states and future outcomes. Such a model could use these relationships to infer missing measurements, predict disease risk and generate possible future health-event trajectories, including their estimated timing.

Large-scale prospective population cohorts offer such an opportunity^14,15^, yet enabling models to naturally learn from cohort data has remained a formidable challenge. A typical large cohort combines demographics, lifestyle exposures, physiological measurements, laboratory biomarkers, molecular profiles, imaging-derived phenotypes and linked longitudinal outcomes^14,16^ for tens to hundreds of thousands of participants^14,15^. These data are predominantly numerical, longitudinal and participant-linked—not in the textual form that language models are designed to process. Machine-learning methods have also been applied to large electronic health record datasets^11,17^. Shared EHR foundation models have also been adapted across hospitals^18^. Crucially, however, LLMs already encode rich biomedical knowledge from their pretraining. We therefore hypothesize that a continued-learning strategy, which enables LLMs to integrate textual biomedical knowledge with prospective cohort data, can substantially enhance model performance across multiple dimensions.

Two recent lines of work show that longitudinal health records can be modelled directly^19^. A generative transformer trained de novo on sequences of coded health events reproduces the rates of more than 1,000 diseases in UK Biobank and transfers to the Danish national registries^20^, and semantic embeddings of medical concepts improve the portability of dedicated risk models across coding systems and healthcare systems^21^. GRASP uses fixed embeddings of medical concepts in a downstream risk model^21^, whereas other work encodes whole patient records and also evaluates model fine-tuning^22^. These studies focus on clinical prediction and do not assess the combination of cohort-based quantitative reasoning and broad medical question answering. We ask whether a medical language model that already encodes biomedical knowledge from text pretraining can additionally absorb quantitative supervision from a prospective cohort without losing the competence with which it began.

Nevertheless, learning from raw cohort data presents several practical challenges. First, tens of thousands of coded clinical fields must be converted into readable sequences^22^ without loss of numerical values, units or the temporal ordering of repeated observations^23^, and the model must learn to interpret these elements. Second, prospective cohort data are participant-centric, requiring the model to understand what each piece of information represents and when it was recorded in order to support prospective reasoning^11,17^. Third, extensive training on cohort data risks eroding the biomedical knowledge already acquired during pretraining, potentially diminishing previously established capabilities^24^.

To address these challenges, we designed a framework for prospective-cohort learning in biomedical LLMs and developed HealthFound as a health world model capable of reasoning over observed health states, inferring missing information and predicting future health events within a unified framework. This framework is specifically designed to enable biomedical LLMs to learn from complex quantitative longitudinal data. Our contributions span data construction, training strategy and evaluation. For data, we converted records from 501,936 UK Biobank participants^14^ into readable longitudinal health profiles through semantic mapping and temporal alignment, supplemented by an additional 7.4 million medical and general reasoning examples to provide broader knowledge and instruction-following supervision. To integrate longitudinal cohort data without eroding existing medical knowledge, we designed a three-stage training framework. A curriculum stage progressively introduces cohort tasks while interleaving medical reasoning data. A rejection-sampled retention stage identifies and rehearses weakened abilities, effectively repairing forgotten knowledge. Finally, a policy optimisation stage uses verifiable rewards to strengthen quantitative reasoning, anchored to the retained model to prevent capability drift^25^. We used multiple benchmarks to evaluate model performance. These include eight public medical benchmarks for assessing general medical competence, as well as tasks for literature-derived question answering, quantitative indicator imputation and future disease prediction across over 1,000 endpoints. We further tested the zero-shot generalizability of HealthFound on two independent datasets—MIMIC-IV and NHANES^26,27^—which represent hospital records and population-survey measurements, respectively. This provides a rigorous assessment of transferability across diverse real-world settings.

## Results

### Study design and evaluation framework

**Figure 1** summarizes the study design, from data construction to evaluation. The cohort data comprised records from 501,936 UK Biobank participants, converted into readable longitudinal health profiles through semantic mapping and temporal alignment. These included approximately 3.8 million disease-prediction examples across 1,074 endpoints, 944,000 natural-language health records, 186,000 quantitative indicator-inference examples and 72,000 scientific question-answer examples. We additionally constructed 7.4 million medical and general reasoning examples. The training mixture totalled approximately 12.4 million examples (**Fig. 1a**).

**Figure 1.**
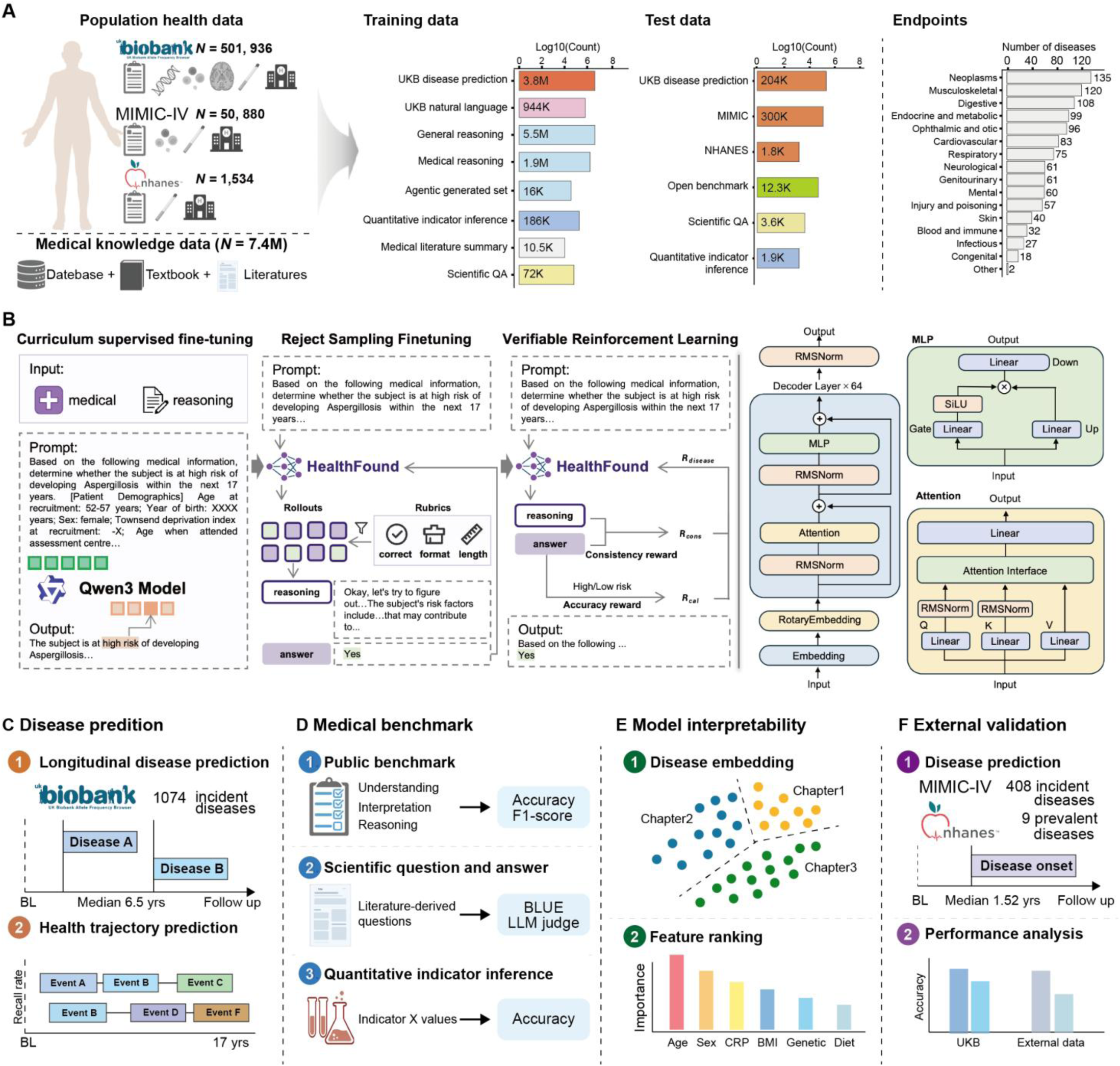
Overview of HealthFound. **a**, Data resources and task construction. UK Biobank records and biomedical knowledge resources provide training and internal evaluation data; MIMIC-IV and NHANES are reserved for external evaluation. The tasks cover disease prediction, natural-language health representation, quantitative indicator inference, scientific question answering and general medical reasoning. The rightmost panel summarizes the distribution of disease endpoints across clinical chapters. **b**, Training framework. HealthFound is initialized from a Qwen3 backbone and trained through curriculum supervised fine-tuning, rejection-sampled retention fine-tuning (RFT), and HealthFound Policy Optimization (HFPO). RFT rehearses medical and general anchor data to maintain medical competence. HFPO applies group-relative policy optimization with task-verifiable rewards, clipped policy updates, reference-policy regularization and an explicit retention loss. The model uses a decoder-only Transformer architecture with rotary positional embeddings, RMS normalization, grouped attention, and gated feed-forward layers. **c**, Longitudinal disease-prediction evaluation across 1,074 incident UK Biobank endpoints and health trajectory prediction. **d**, Evaluation on public medical benchmarks, literature-derived scientific questions, and quantitative indicator inference. **e**, Model-interpretability analyses based on disease representations and the contribution of health-feature groups to model reasoning. **f**, External evaluation on 408 incident disease tasks in MIMIC-IV and nine prevalent disease tasks in NHANES. BL denotes baseline and F1 denotes F1 score.

HealthFound was trained through three stages (**Fig. 1b**). First, curriculum supervised fine-tuning (SFT) gradually increased the proportion of cohort-based training examples while retaining medical and general reasoning data, enabling the model to learn from longitudinal health records. Second, rejection-sampled retention fine-tuning (RFT) generated candidate responses to disease prediction task, selected those passing correctness and formatting checks, and used them alongside the original answers to preserve the model’s existing reasoning abilities. Finally, HealthFound Policy Optimization (HFPO) improved quantitative reasoning ability using task-specific rewards for answer accuracy and consistency, while limiting divergence from the RFT model and continuing to reinforce general reasoning abilities.

We evaluated HealthFound across several downstream tasks (**Fig. 1c–f**). Using UK Biobank data, we assessed disease prediction across 1,074 endpoints using only pre-onset health records, generation of future health-event trajectories, and inference of missing biomedical measurements. We further evaluated medical knowledge and reasoning using eight public medical benchmarks and literature-derived question answering. To explore what the model had learned, we compared disease representations with those of the unadapted backbone and examined how masking clinical features affected prediction accuracy. Finally, we assessed zero-shot generalization on two external datasets: MIMIC-IV (408 incident disease endpoints) and NHANES (nine prevalent-disease tasks).

### Predicting future disease onset across 1,074 endpoints

We evaluated disease classification across 1,074 UK Biobank endpoints^14,15^ using health records truncated before the first recorded diagnosis (**Fig. 2**). The interval from the end of the input record to diagnosis had a median of 6.5 years and a maximum of 16.9 years. All models received the same health profiles and returned high-risk or low-risk predictions. Accuracy, positive-class F1, sensitivity and specificity were evaluated for each endpoint.

**Figure 2.**
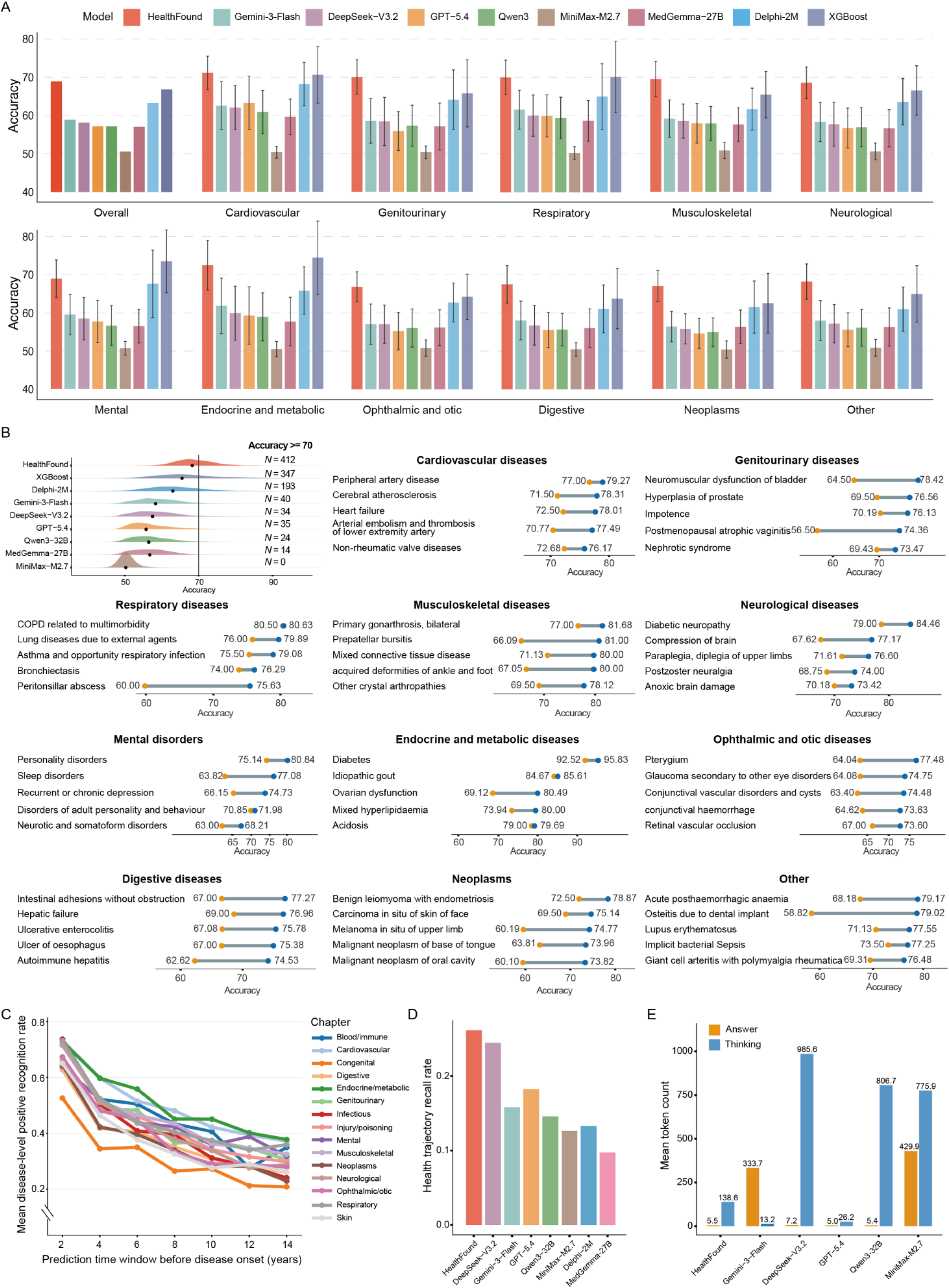
Population-scale disease prediction in UK Biobank. **a**, Disease-level prediction accuracy summarized by clinical chapter for HealthFound, the comparator language models and traditional machine learning models. Bars show the mean disease-level accuracy within each chapter and error bars denote the mean ± 1 s.d. across the endpoints contributing to that chapter. The first group reports performance across all evaluated endpoints. **b**, Distribution and endpoint-level analysis of prediction performance. The ridgeline plot shows the distribution of disease-level accuracy for each model and the number of endpoints achieving an accuracy of at least 70%. Dumbbell plots compare HealthFound with GPT-5.4 on representative endpoints from major disease chapters. **c**, Mean chapter-level positive-recognition rate as a function of the interval between the input cut-off and first disease onset. The positive-recognition rate is sensitivity among cases. Each line represents one clinical chapter, and the vertical axis is truncated below the plotted range. Points represent consecutive 2-year intervals, defined as left-closed and right-open, and are plotted at their upper boundaries. **d,** Health-event trajectory recall for different models. **e,** Mean number of generated tokens in the reasoning and final-answer fields for each model. Token counts are descriptive because access to internal reasoning tokens differs across model interfaces.

Compared with general-purpose LLMs, HealthFound achieved the highest mean accuracy in every clinical chapter (**Fig. 2a**). Its overall mean accuracy was approximately 68.9%. Relative to the strongest comparator within most chapters, the mean advantage was 10.0 percentage points and ranged from 7.9 to 11.5 percentage points. This advantage was observed across endocrine/metabolic, cardiovascular, genitourinary, respiratory, musculoskeletal, mental, neurological, digestive, ophthalmic and neoplastic disease groups.

A total of 412 of the 1,074 endpoints reached an accuracy of at least 70% with HealthFound, compared with 347 for XGBoost, 193 for Delphi-2M, 40 for Gemini-3-Flash, 34 for DeepSeek-V3.2, 35 for GPT-5.4, 24 for Qwen3, 14 for MedGemma-27B, and none for MiniMax-M2.7. In representative comparisons with GPT-5.4, accuracy was 78.01% versus 72.50% for heart failure and 84.46% versus 79.00% for diabetic neuropathy (**Fig. 2b**). Differences were also observed among respiratory, genitourinary, neurological and ophthalmic endpoints, consistent with the broad advantage observed across clinical chapters.

Comparisons with publicly available medical LLMs tested whether this advantage extended to domain-specialized models. HealthFound achieved the highest overall disease-prediction accuracy, exceeding the next-best medical LLM, MedGemma-27B, by 11.8 percentage points (**Fig. 2a** and **Supplementary Table 1**). Comparisons with Delphi-2M, XGBoost and logistic regression also favoured HealthFound, with an advantage of 5.9, 2.3 and 7.3 percentage points (**Fig. 2a** and **Supplementary Table 2**). We further evaluated 23 low-prevalence diseases at observed case proportions of 0.05%–5%. HealthFound retained the highest overall performance in this comparison. (**Supplementary Fig. 1** and **Supplementary Fig. 2)**

Chapter-level positive-case detection rates were generally lower among cases with longer intervals between the input cut-off and the first recorded diagnosis (**Fig. 2c**). Across clinical chapters, rates were approximately 0.50–0.74 in the 0–2-year interval, compared with 0.20–0.40 in the 12–14-year interval. **Fig. 2d** summarizes trajectory recall across models. HealthFound used a mean of 138.6 reasoning tokens per prediction, fewer than DeepSeek-V3.2 (985.6), Qwen3 (806.7) and MiniMax-M2.7 (775.9), but more than GPT-5.4 and Gemini-3-Flash (**Fig. 2e**). The performance advantage was therefore not associated with longer generated reasoning traces.

### Predicting longitudinal health-event trajectories

To evaluate whether HealthFound could predict how an individual’s health might evolve over time, we compared it with Delphi-2M and other LLMs on longitudinal health-event generation (**Fig. 2d**). We selected 10,000 UK Biobank participants and used their health records before a defined prediction cut-off to generate future trajectories. Each trajectory consisted of predicted disease events and their estimated occurrence times. The evaluation included 77,564 recorded events after the cut-off, covering newly occurring inpatient diagnoses and death over a maximum follow-up window of 17 years. We evaluated each model using eight valid trajectories per participant, matching the sampling count reported for HealthFlux^28^. We assessed performance using the recall of event recovery, which measured the proportion recovered in at least one of the eight trajectories. HealthFound achieved a recall of 26.1%, compared to 13.3% for Delphi-2M and 24.5% for DeepSeek-V3.2 (Fig. 2d).

### Inferring missing quantitative indicators from health profiles

We evaluated whether HealthFound could infer missing quantitative measurements from the remaining information in a health profile. Using approximately 1,900 held-out examples, we removed one observed measurement from each profile and asked the models to predict its value (**Fig. 3a**). Prediction accuracy was evaluated at relative-error tolerances ranging from 0.1% to 5%.

**Figure 3.**
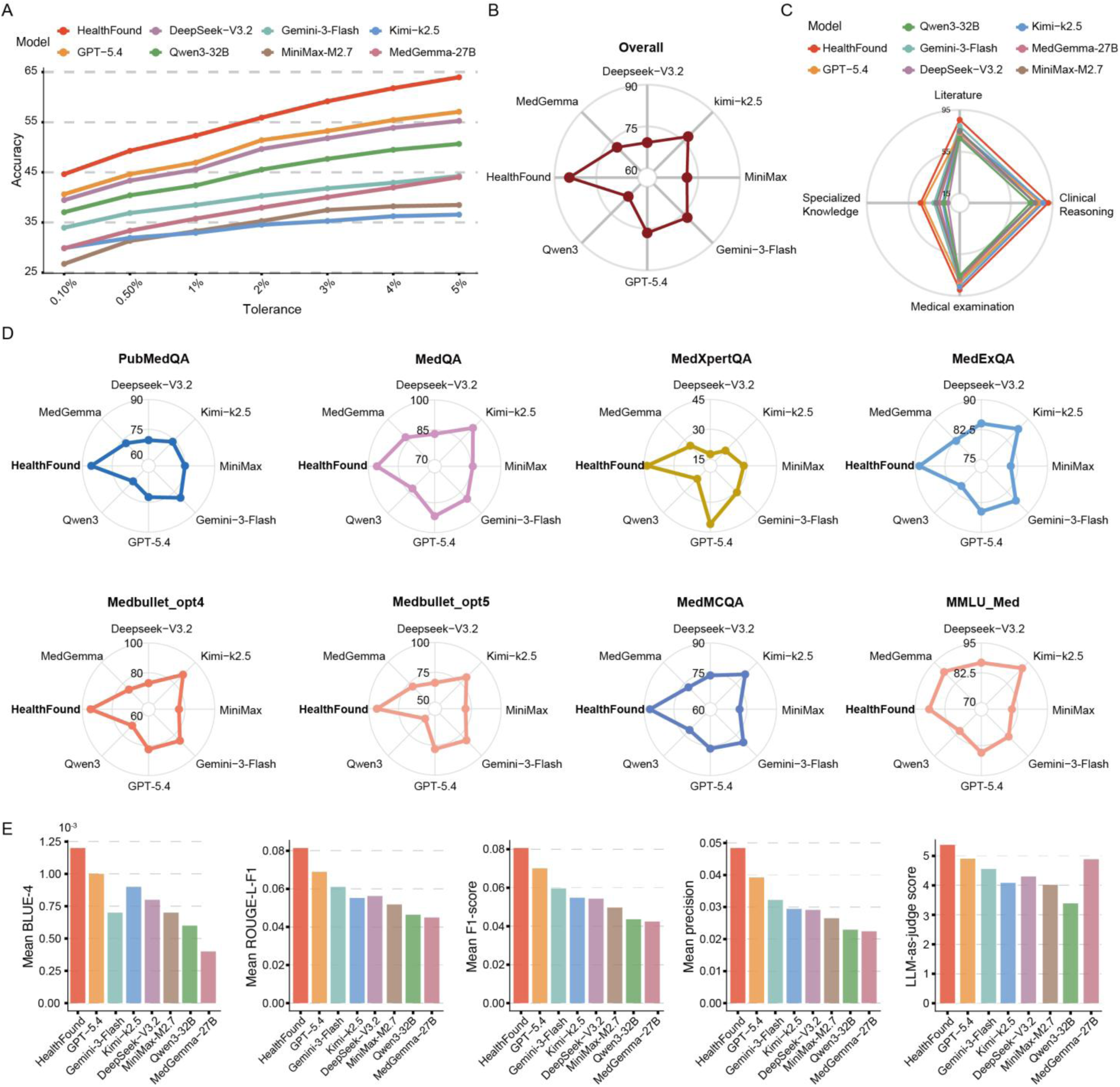
Quantitative indicator inference and evaluation on medical and scientific reasoning benchmarks. **a**, Quantitative indicator-inference accuracy. Models recover masked physiological or laboratory values from the surrounding health profile; a prediction is counted as correct when its relative numerical error falls within the indicated tolerance from 0.1% to 5%. **b**, Mean accuracy across eight established public medical benchmarks. **c**, Performance grouped into literature comprehension, clinical reasoning, medical examination, and specialized medical knowledge. **d**, Model-level comparisons on PubMedQA, MedQA, MedXpertQA, MedExQA, Medbullets-opt4, Medbullets-opt5, MedMCQA, and the medical subset of MMLU. Radial axes show benchmark accuracy. **e,** Scientific question-answering evaluation using BLEU-4, ROUGE-L F1, word-overlap F1, precision and LLM-as-a-judge scores.

HealthFound consistently achieved the highest accuracy among the evaluated language models across all tolerance thresholds. At the primary 3% tolerance, HealthFound achieved an accuracy of 59.16%, compared with 53.23% for GPT-5.4, an improvement of 5.93 percentage points. At the stricter 1% tolerance, accuracies were 52.32% and 46.93%, respectively. At the 5% tolerance, they increased to 63.96% and 57.07%, corresponding to a 5.66-percentage-point advantage for HealthFound (**Supplementary Table 3**).

We further compared HealthFound with conventional statistical approaches, including mean imputation and multiple imputation by chained equations (MICE). HealthFound achieved substantially higher accuracy than both methods across the evaluated tolerance thresholds (**Supplementary Fig. 3**). Together, these results demonstrate HealthFound’s ability to infer missing quantitative measurements from complex health profiles, while highlighting differences in performance across indicator types.

### Retention of general medical competence on public benchmarks

To assess medical competence after cohort specialization, HealthFound was evaluated on eight independent public benchmarks spanning medical examinations, clinical reasoning, literature comprehension and specialized medical knowledge (**Fig. 3b–d**).

HealthFound reached a mean accuracy of 84.70%, 7.23 percentage points above the next-best aggregate result of 77.47% from Kimi-k2.5 (**Fig. 3b**). The model ranked first on seven of the eight individual benchmarks. It scored 95.76% on MedQA, 94.48% on Medbullets-opt4, 93.18% on Medbullets-opt5, 88.94% on MedExQA and 43.55% on MedXpertQA; on MMLU-Med, Kimi-k2.5 scored 91.50% compared with 89.10% for HealthFound (**Fig. 3d**). These results show that quantitative specialization was compatible with broad medical benchmark performance.

### Training stages support complementary capabilities

The sequential-stage evaluation demonstrated distinct contributions from each training stage (**Supplementary Table 4**). Curriculum supervised fine-tuning (SFT) increased UK Biobank disease-prediction accuracy from 57.1% to 61.9% and mean medical-benchmark accuracy from 66.3% to 80.0%, indicating improvements in both cohort-based prediction and medical reasoning. Rejection-sampled retention fine-tuning (RFT) further improved medical-benchmark accuracy to 84.3%, while UK Biobank accuracy increased to 62.8%. Finally, HealthFound Policy Optimization (HFPO) substantially improved disease-prediction accuracy to 68.9%, while maintaining medical-benchmark accuracy at 84.7%. Across the three stages, quantitative indicator-inference accuracy also increased progressively from 37.1% to 49.3%, while ROUGE-L F1 for literature-derived question answering improved from 0.0464 to 0.0814. These results suggest that the three stages contributed complementary improvements in quantitative prediction and medical reasoning.

To examine whether these gains resulted from increased training-data volume alone, we compared alternative configurations using matched data volumes (**Supplementary Table 4**). Standard UK Biobank SFT achieved 58.1% disease-prediction accuracy, while conventional SFT using data from all three stages reached 60.0%. Applying conventional GRPO after sequential SFT and RFT reduced prediction accuracy on UKB from 62.8% to 61.4%, suggesting that standard GRPO is not well suited to our disease-prediction task and may even degrade predictive performance, whereas our proposed HFPO provides a more effective optimization strategy. In addition, we compared SFT followed by Reply with SFT followed by RFT. The inferior performance of the former supports the effectiveness of our RFT design. These findings suggest that the sequential training strategy and task-specific optimization contributed to performance improvements beyond those attributable to training-data volume alone.

### Literature-derived question answering

We evaluated answers to 3,574 held-out questions derived from biomedical publications. The questions covered basic and clinical medicine, biostatistics, bioinformatics and medical research methods. Each model answered the same questions under a common prompt, and its responses were compared with reference answers using BLEU, ROUGE and token-level precision. These measures quantify agreement with the reference wording rather than clinical correctness. Among the models shown in Fig. 3e, HealthFound achieved the highest mean scores on the four displayed lexical-overlap metrics, including mean ROUGE-L F1 of approximately 0.081, token-level F1 of 0.081, precision of 0.048 and BLEU-4 of approximately 0.001 (**Fig. 3e**).

Since lexical-overlap metrics do not directly assess scientific correctness, we therefore evaluated answer correctness separately using three independent LLM judges. HealthFound received the highest mean answer-correctness score (5.4/10, **Supplementary Table 5**), exceeding the next-best model by approximately 9.25% and supporting stronger agreement with reference scientific conclusions.

### Disease representations reveal clinically coherent organization

We examined the clinical organization of disease representations by comparing HealthFound with the Qwen3-32B backbone. Clinically related diseases clustered in the representation space (**Figure 4a**), with clearer separation and more coherent clinical organization in HealthFound than in the Qwen3-32B backbone (**Supplementary Fig. 4**).

**Figure 4.**
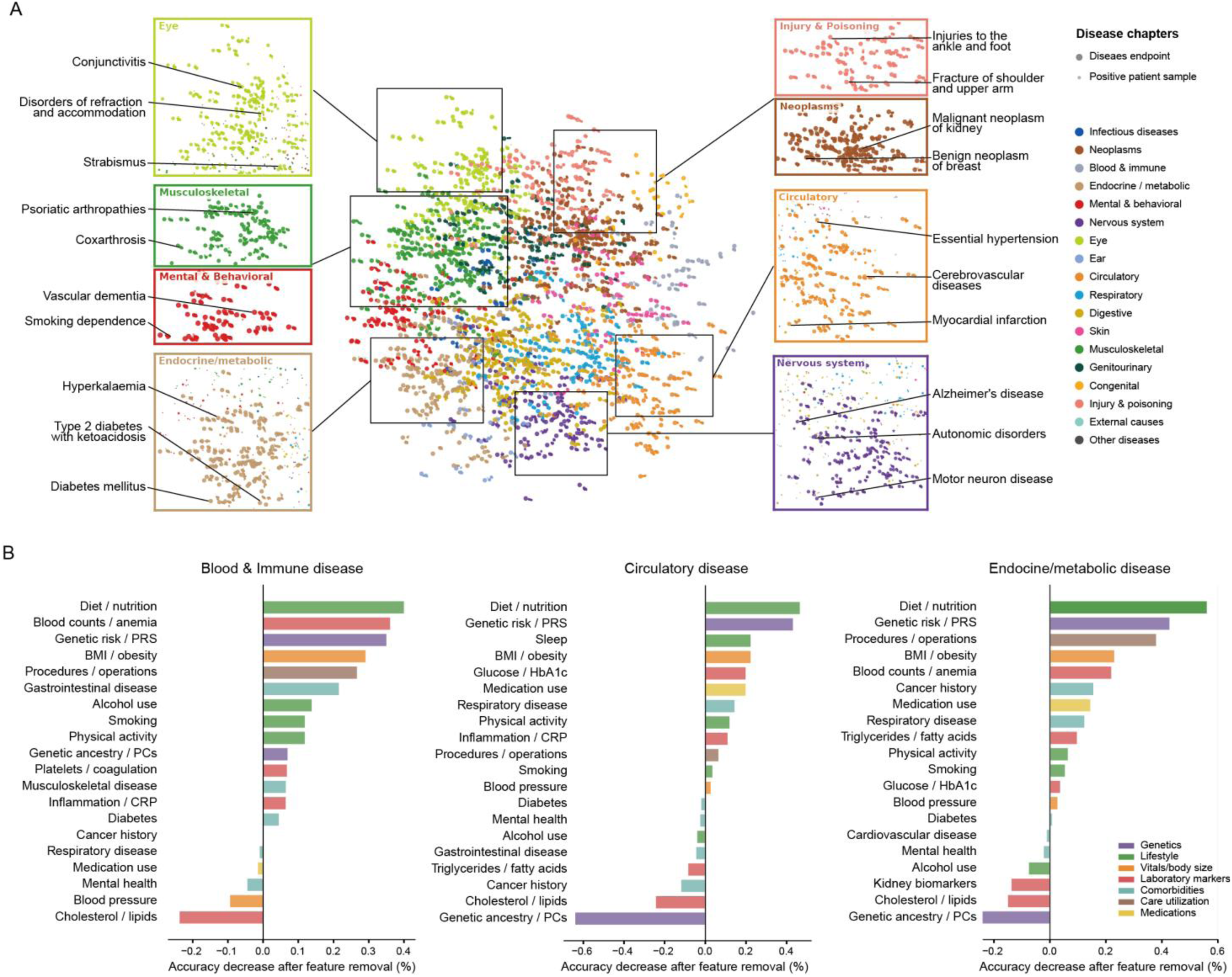
Structure of learned disease representations and feature importance. **a**, Two-dimensional projection of representations for disease-prediction endpoints. Colours denote clinical chapters and dots mark selected labelled endpoints. Insets show selected disease groups and labelled endpoints. **b,** Changes in prediction accuracy after individual feature groups were removed. Examples shown here are blood and immune, circulatory and endocrine/metabolic diseases.

Input-masking experiments further showed that removing key biomedical variables reduced disease-prediction accuracy, supporting the predictive relevance of features identified from the model’s reasoning. Overall, after removing the top-ranked PRS and diet/nutrition variables for blood and immune diseases, circulatory diseases and endocrine/metabolic diseases, prediction accuracy decreased by more than 0.3%. In particular, removing the top-ranked blood count and anemia variables for blood and immune diseases reduced prediction accuracy by approximately 0.35%, while removing the sleep variable for circulatory diseases resulted in an approximately 0.2% reduction (**Figure 4b**).

### Qualitative clinical examples

We examined how HealthFound combined temporal and clinical information in individual predictions. Three representative cases illustrated prediction before disease onset, reasoning over a longitudinal record and treatment selection (**Fig. 5**). HealthFound and Gemini-3-Flash received the same record and question, and the comparison highlights the information emphasized in their responses.

**Figure 5.**
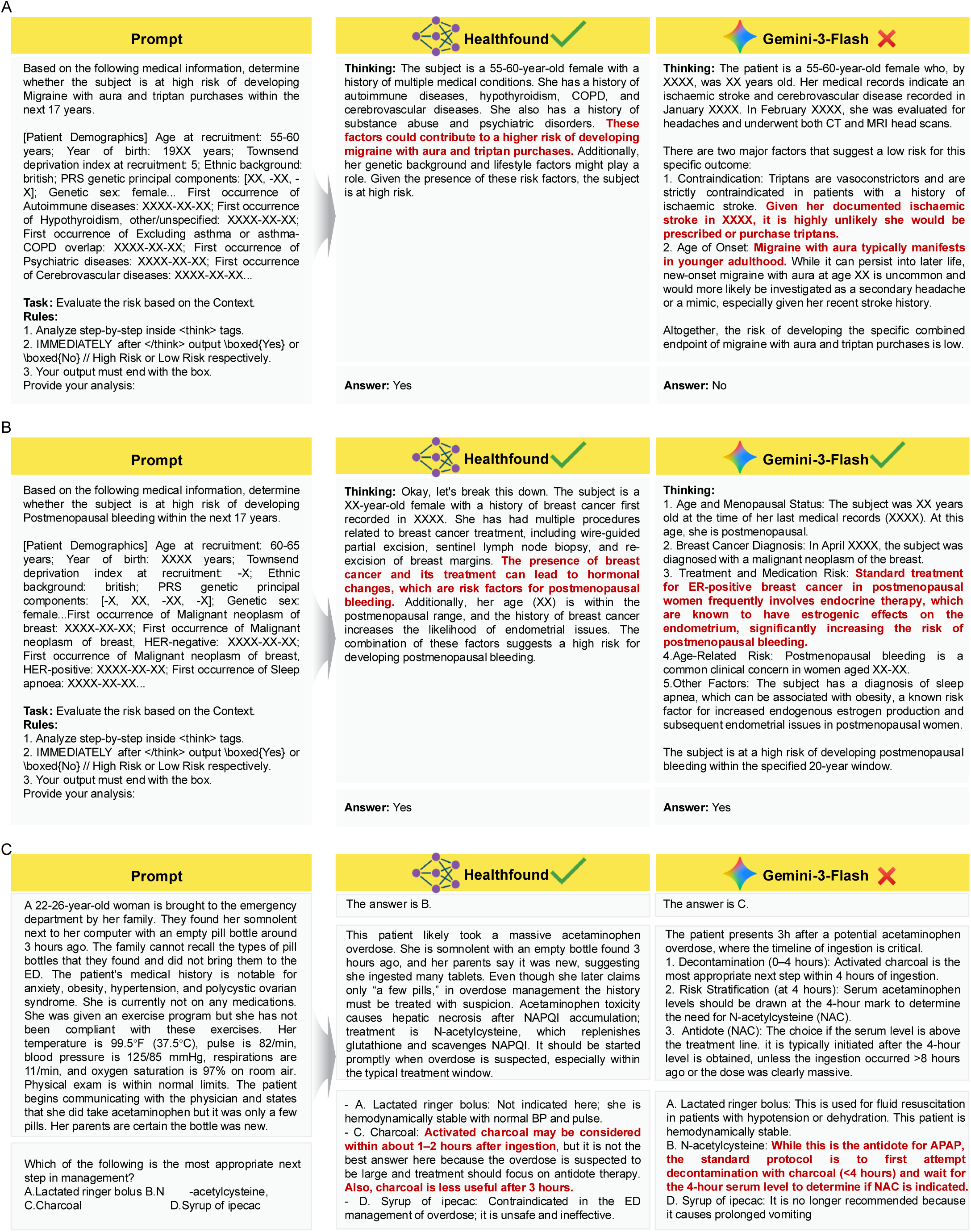
Representative qualitative comparisons of clinical reasoning. **a**, Prediction of future hydrocephalus from a longitudinal health record. HealthFound identifies the high-risk label, whereas Gemini-3-Flash returns a low-risk prediction. **b**, Prediction of postmenopausal bleeding in a participant with a history of breast cancer. Both models return the reference high-risk label, while emphasizing partly different evidence. **c**, Medical examination question concerning management after a suspected acetaminophen overdose. HealthFound selects N-acetylcysteine and explains the time-sensitive treatment rationale, whereas Gemini-3-Flash selects activated charcoal. Red text highlights selected statements in the generated explanations. Check marks and crosses indicate agreement and disagreement with the final reference answer.

In the hydrocephalus example, HealthFound predicted the reference high-risk label, whereas Gemini-3-Flash predicted low risk (**Fig. 5a**). Both models produced the reference high-risk label in the postmenopausal-bleeding example, and both models considered the history of breast cancer and its treatment (**Fig. 5b**). In the acetaminophen-overdose question, HealthFound selected N-acetylcysteine and described the time-sensitive antidote rationale, whereas Gemini-3-Flash selected activated charcoal (**Fig. 5c**).

To examine the contribution of temporal information, we perturbed the dates of existing clinical events while preserving their original chronological order and retaining the biomedical measurements. Disease-prediction accuracy decreased, with many previously correct predictions becoming incorrect. Some predictions remained correct after perturbation, consistent with temporal information contributing alongside other biomedical risk indicators (**Supplementary Table 6**).

### Zero-shot transfer to independent datasets

We evaluated zero-shot transfer of HealthFound to two independent datasets (**Fig. 6**). MIMIC-IV provided longitudinal hospitalization records from 50,880 individuals and 408 incident disease endpoints; inputs were restricted to information recorded before the target diagnosis. NHANES provided cross-sectional examination and laboratory measurements from 1,534 survey participants for nine prevalent-disease tasks. HealthFound was evaluated without further training, reward optimization or prompt adjustment.

**Figure 6.**
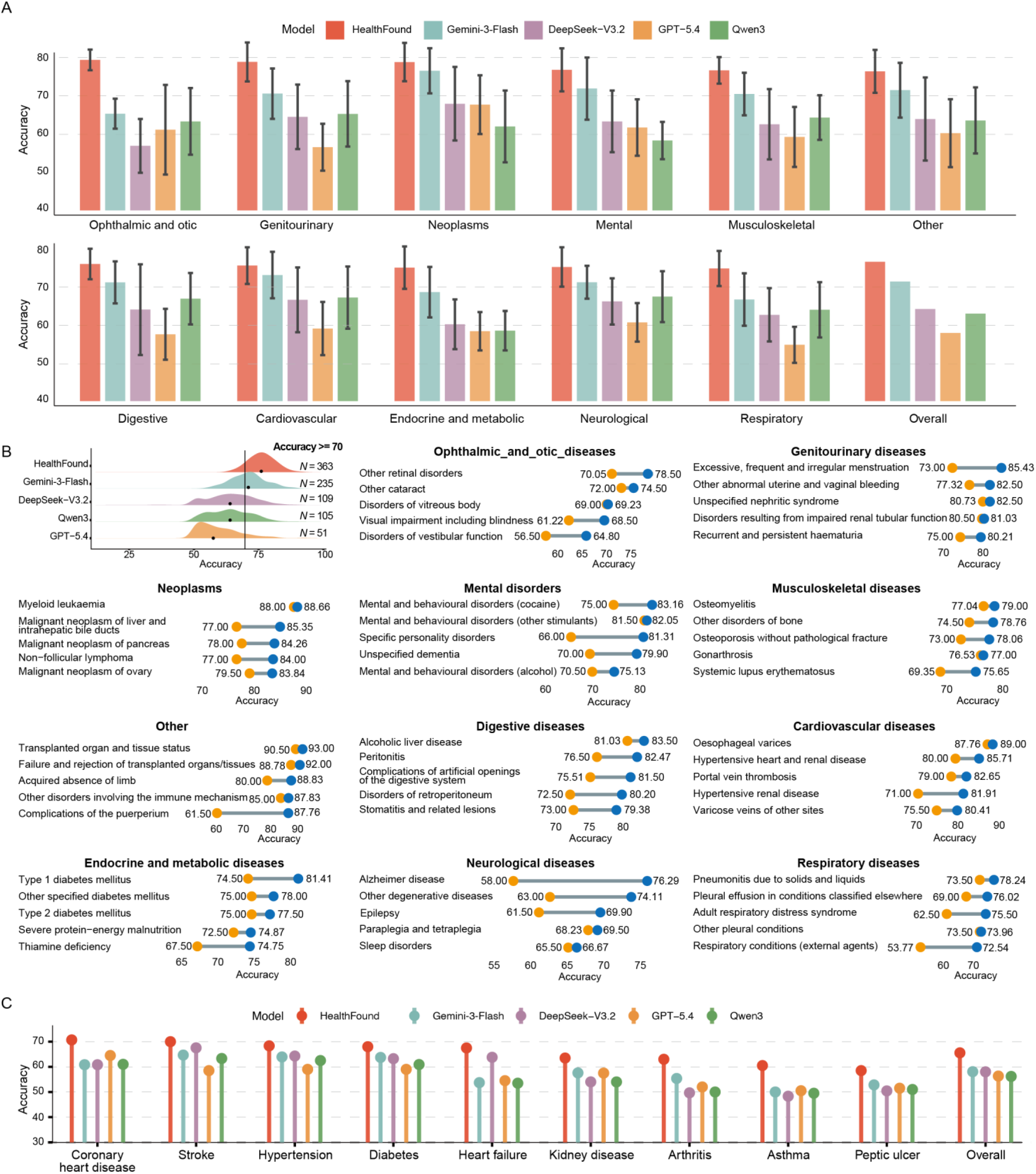
External evaluation of disease-prediction performance. **a**, Accuracy across clinical chapters for 408 incident disease endpoints constructed from MIMIC-IV. Bars show the mean disease-level accuracy within each chapter and error bars denote the mean ± 1 s.d. across the endpoints contributing to that chapter. The final group summarizes all evaluated MIMIC-IV endpoints. **b**, Distribution and endpoint-level analysis of MIMIC-IV performance. The ridgeline plot shows the distribution of endpoint-level accuracy and the number of endpoints reaching at least 70% accuracy. Dumbbell plots compare HealthFound with GPT-5.4 on representative endpoints from each clinical chapter. **c**, Accuracy on nine prevalent-disease classification tasks constructed from NHANES, including coronary heart disease, stroke, hypertension, diabetes, heart failure, kidney disease, arthritis, asthma and peptic ulcer. Points show model accuracy for each endpoint and for the aggregate NHANES comparison.

On 408 MIMIC-IV incident endpoints, HealthFound achieved the highest mean accuracy in every disease chapter and an overall accuracy of 76.1%. Gemini-3-Flash reached approximately 71.0%, and the remaining general-purpose comparators reached 63.9% or lower (**Fig. 6a**). A total of 363 endpoints reached at least 70% accuracy with HealthFound, compared with 235 for Gemini-3-Flash, 109 for DeepSeek-V3.2, 105 for Qwen3 and 51 for GPT-5.4 (**Fig. 6b**). Representative comparisons with GPT-5.4 included 81.9% versus 71.0% for hypertensive renal disease, 76.3% versus 58.0% for Alzheimer disease and 72.5% versus 53.8% for respiratory conditions due to external agents. **Supplementary Fig. 5** shows MIMIC-IV performance by prevalence stratum.

On the nine NHANES prevalent-disease tasks, HealthFound achieved the highest accuracy for every reported endpoint and a mean accuracy of 65.6% overall (**Fig. 6c**), compared with 58.0% for Gemini-3-Flash and DeepSeek-V3.2, and lower than 56.4% for the remaining general-purpose comparators. The tasks covered coronary heart disease, stroke, hypertension, diabetes, heart failure, kidney disease, arthritis, asthma and peptic ulcer. Performance was strongest on coronary heart disease and stroke and was lower for arthritis, asthma and peptic ulcer, but HealthFound remained the highest-performing model across these tasks.

## Discussion

HealthFound demonstrates that longitudinal population cohorts can extend medical language models beyond medical knowledge retrieval towards quantitative reasoning over evolving health states while retaining broad medical competence. Whereas biomedical literature, clinical vignettes and examination questions primarily describe established diseases and their management^3,4,10^, prospective cohorts link measurements from apparently healthy or preclinical individuals to health outcomes observed years later^14,15^. By integrating these complementary sources of information through a three-stage training framework, HealthFound supports basic LLM functions together with incident disease prediction, missing-measurement inference, future health-event trajectory generation and scientific question answering. Its advantages also extended to external datasets, demonstrating zero-shot transfer across different healthcare settings and data structures.

HealthFound differs from recent cohort- and record-based predictors in how longitudinal cohort information is incorporated into the model. Delphi-2M is trained from scratch on sequences of coded health events^20^, whereas GRASP uses an LLM to embed medical concepts before training a separate transformer for disease-risk prediction^21^. HealthFound instead updates a pretrained language model through cohort-derived supervision. The resulting model predicts 1,074 incident disease endpoints, infers missing biomedical measurements and answers scientific questions while achieving 84.70% mean accuracy across eight public medical benchmarks. Its ability to generate future health-event trajectories further extends these capabilities. HealthFound also outperformed Delphi-2M, XGBoost and logistic regression in incident disease prediction. These findings provide a practical basis for health world modelling, in which a single language model uses longitudinal health information to infer missing measurements, estimate disease risk and generate possible future health events.

Maintaining broad medical competence was an important part of this training strategy. Curriculum SFT introduced cohort-based tasks while retaining medical and general reasoning examples. RFT subsequently reinforced these abilities, increasing mean accuracy across eight public medical benchmarks while producing only a modest improvement in incident disease prediction. HFPO then increased disease-prediction accuracy while maintaining medical-benchmark performance. Moreover, conventional SFT using the combined training data reached 60.0% UK Biobank accuracy, compared with 68.9% for the complete framework. The improvement therefore depended on how the model was trained, beyond the amount of training data alone.

The representation analyses provide further evidence that cohort training changed how HealthFound organized disease-related information. Although clinical chapter labels were not included in the embedding inputs, clinically related diseases formed clearer groups in HealthFound than in the Qwen3-32B backbone. Input-masking experiments also showed that removing selected biomedical variables reduced disease-prediction accuracy, consistent with the clinical features identified in the model’s generated reasoning. These findings suggest that HealthFound learned useful relationships among diseases and biomedical measurements.

External evaluations further tested whether these capabilities extended beyond UK Biobank. Without additional training or prompt adjustment, HealthFound ranked the top in mean accuracy across 408 MIMIC-IV endpoints and nine NHANES endpoints. These results extend HealthFound’s performance to hospital records and population-survey data with different observation patterns and disease definitions, supporting its ability to apply cohort-derived quantitative reasoning beyond the original UK Biobank setting.

Future work has three priorities. First, evaluation in more diverse longitudinal cohorts is needed to assess generalizability across populations and healthcare settings, including the effects of selection bias^29^, diagnostic coding and differences in follow-up. Second, richer modelling of health trajectories could better capture repeated measurements, irregular observation times, informative missingness and multimodal data beyond text-serialized derived features. Third, prospective studies should evaluate risk calibration under natural disease prevalence, the accuracy of temporal reasoning and explanations, and whether model predictions can improve clinical decision-making and patient outcomes^30^.

In conclusion, HealthFound shows that longitudinal population data can train medical language models to analyse quantitative health profiles while retaining broad medical competence. By integrating cohort-derived longitudinal information with pretrained medical knowledge, HealthFound unifies disease prediction, missing-measurement inference, future health-event trajectory generation and scientific question answering within a single model. Its performance across thousands of endpoints, established medical benchmarks and two external datasets points towards a health world model that moves beyond static medical knowledge to reason quantitatively about evolving human health states and their possible future outcomes.

## Data availability

All data used in this study were accessed and analysed in compliance with the applicable data use agreements and policies of the respective cohorts and biobanks. UK Biobank received approval from the National Information Governance Board for Health and Social Care and NHS North West Multi-centre Research Ethics Committee (11/NW/0382). Participants gave written informed consent. Individual-level UK Biobank data are available to approved researchers through the UK Biobank access procedure and cannot be redistributed by the authors. MIMIC-IV v3.1 is available through PhysioNet subject to its credentialing and data-use requirements. NHANES 1999–2000 data are publicly available from the US Centers for Disease Control and Prevention. Training also used the Medical-O1-Reasoning series, GlaiveAI Reasoning-v1-20m and tulu-3-sft-personas datasets, together with the biomedical literature resources described in Methods. Public medical benchmarks are listed in **Supplementary Table 7**. Dataset access and licensing information is provided in **Supplementary Table 8**.

## Code availability

The project repository will be available at github.

## Acknowledgements

This study used the UK Biobank Resource under application numbers 202239 and 19542. We thank all participants and team members of the UK Biobank. This study was supported by the Noncommunicable Chronic Diseases–National Science and Technology Major Project (2025ZD0546300) and the 2025 Yangtze River Delta Science and Technology Innovation Community Joint Research (Basic Research) Project (2025CSJZN00100).

## Author contributions

H.G., L.S., P.R., J.H. and Y.W. contributed equally to this study. L.S. implemented the models and performed three stages of model training. J.H. designed and formalized the training tasks and constructed the corresponding datasets. Y.W. conducted comprehensive evaluations of model performance across the tasks in this study. P.R. prepared the visualizations of the results. L.S., P.R., J.H. and Y.W. contributed to writing the manuscript. H.G., J.F., W.G. and W.C. conceived the study, designed the overall research framework, jointly supervised the research and led the writing and revision of the manuscript. L.S., J.H., Y.W., H.L., J.D. and J.W. contributed to code development. All authors discussed the results, reviewed the manuscript and approved the final version.

## Methods

### Natural-language training data construction

To express heterogeneous measurements in a common input format, we converted cohort records to text before constructing task-specific examples.

UK Biobank provided data from 501,936 participants recruited at ages 40–69 years in 2006–2010^14,15^. Measurements included demographics, lifestyle and diet, physical measurements, blood biochemistry and haematology, polygenic risk scores (PRSs) (**Supplementary Table 9**), imaging-derived phenotypes and linked clinical records. Median cohort follow-up was 14.1 years, with a maximum of approximately 17 years. UK Biobank contributed approximately 3.8 million disease-prediction examples covering 1,074 endpoints, 944,000 natural-language health records and 186,000 masked-measurement examples.

We first converted the structured UK Biobank records into readable health profiles by mapping field identifiers to clinical names, data types, coding hierarchies and units. Missing values and ambiguous codes were omitted, while categorical codes were translated into their documented meanings and numerical values were retained with their units and assessment dates. These standardized profiles were then used to construct four types of training examples.

#### Disease-prediction examples

For disease prediction, we identified incident cases and eligible controls for each of the 1,074 disease endpoints and sampled the two groups in balanced numbers. Participants with a pre-existing diagnosis of the target disease were excluded. Each input contained only observations available before the prediction cut-off, without summarization or information recorded after that point. To accommodate lengthy records, we divided the profiles into segments of up to 4,096 tokens, with an overlap of 300 tokens between adjacent segments. Each segment began with sex and age at recruitment and was paired with the corresponding endpoint-specific label. This process generated approximately 3.8 million disease-prediction examples, including multiple endpoint-specific examples and record segments from individual participants.

#### Natural-language health records

To construct the 944,000 natural-language health-record examples, we instead divided the standardized observations into text blocks and used a locally deployed Qwen3 model to summarize each block. The summaries were subsequently merged into coherent health reports that grouped clinically related information and consolidated repeated descriptions while preserving measurement values, units, assessment dates and follow-up findings. Summarization was performed using vLLM^31^ with temperature 0.6 and top-p 0.9.

#### Masked-measurement examples

We constructed 186,000 masked-measurement examples by removing one observed measurement from a health profile and asking the model to infer it from the remaining information. The removed measurement served as the reference answer. These examples covered different indicator domains, including haematology, anthropometry, biochemistry and polygenic risk scores, with the expected answer format determined by the type of measurement.

#### Agent-generated task examples

To extend cohort-derived supervision beyond predefined disease-prediction and measurement-inference tasks, we used an agent pipeline to generate 15,552 additional instructions from UK Biobank health profiles. The pipeline covered six task types: risk stratification, disease prediction, prognosis, estimation of missing-measurement ranges, lifestyle intervention and comorbidity. For each candidate, the agent generated a structured output containing the instruction, input health profile, reasoning, conclusion, task type and confidence. Its memory retained task specifications, operating rules, relevant context and previous validation errors to guide subsequent generation. We then applied structural and content checks to ensure that all required fields were present, reasoning and conclusions were relevant to the assigned task, and each generation batch covered all six task types. For disease-prediction tasks, candidates were rejected if the input revealed the target diagnosis or included observations after the prediction cut-off, or if the reasoning used an already confirmed diagnosis to assign risk. Candidates that failed validation were allowed one targeted revision using the original health record and the identified errors. Only candidates that passed the complete validation procedure were retained for training.

In addition to UK Biobank examples, the training corpus also contained 72,000 scientific question–answer pairs and 7.4 million medical and general reasoning examples (**Supplementary Table 10**).

#### Medical and general reasoning examples

To preserve the model’s general reasoning and medical knowledge during cohort-specific training, we assembled 5.5 million general reasoning examples and 1.9 million medical reasoning examples. The general reasoning data included examples from GlaiveAI Reasoning-v1-20m, covering logical reasoning, multistep problem solving and instruction following. Medical reasoning examples covered diagnosis, differential diagnosis, risk-factor identification, test selection, treatment recommendations and clinical decision-making. Sources included Medical-O1-Reasoning^32^ and tulu-3-sft-personas^33^. We converted the examples into a common Alpaca-style instruction format, retaining the question, reference answer and reasoning trace when available. After standardizing the fields and removing duplicate records and unnecessary special characters, we randomly subsampled the datasets to control their relative contributions to training. This process produced 7.4 million medical and general reasoning examples.

#### Scientific question–answer pairs

To train the model to interpret biomedical research evidence, we constructed scientific question–answer pairs from 10,500 curated publications based on UK Biobank. We used DeepSeek-V3.2 to process the article texts and extract structured study information, including research questions, data sources, sample sizes, variable definitions, statistical methods, quantitative findings and main results. We separately extracted the authors’ interpretations, including proposed mechanisms, clinical implications and study limitations. We then paired the central research questions with their corresponding conclusions to construct question–answer examples grounded in the extracted evidence. This procedure generated 75,574 question–answer pairs, of which 72,000 were used for training and 3,574 were reserved for evaluation.

### Model architecture and training

HealthFound was built on Qwen3-32B, a decoder-only language model. We retained its original architecture, tokenizer and vocabulary of 152,064 tokens, without adding task-specific prediction heads. All inputs, including instructions, health measurements and numerical values, were represented as text. Training sequences were limited to 4,096 tokens, while the inference context window was configured separately. We adapted the model through three successive training stages, each serving a different purpose.

#### Curriculum supervised fine-tuning (SFT)

We first trained the model to generate reference answers for cohort-derived, medical and general reasoning tasks. During training, we gradually increased the proportion of cohort examples while continuing to include medical and general reasoning data. This stage introduced longitudinal health-profile reasoning while maintaining exposure to the model’s existing knowledge and reasoning tasks.

#### Rejection-sampled retention fine-tuning (RFT)

We next reinforced previously learned medical and general reasoning abilities. The model generated candidate responses to selected anchor prompts, and responses were retained if they passed correctness and formatting checks. We then combined the accepted responses with the original reference answers for further supervised training. The resulting model was used to initialize the final training stage and served as its fixed reference model.

#### HealthFound Policy Optimization (HFPO)

Finally, we improved task-specific performance by rewarding correct and consistent responses. For each prompt, the model generated multiple candidate answers, which were scored using task-specific rules. In disease prediction, the reward primarily reflected final-answer correctness (weight 0.8), with additional contributions from reasoning-label correctness (0.1) and consistency between the reasoning and final answer (0.1). The model was updated to favour higher-scoring responses within each group, while limiting its divergence from the RFT reference model and retaining supervised training on anchor answers. The number of training examples used at each stage is reported in **Supplementary Table 10**. Task-specific reward components and their weights are listed in **Supplementary Table 11**.

### Evaluation protocol and comparators

All language models were evaluated using the same task-specific prompts, input health records and required answer formats. Where supported by the model interface, we used a temperature of 0.6, top-p of 0.9 and a maximum of 16,384 generated tokens. Depending on the task, we extracted the final disease-risk label, numerical prediction or written answer and compared it with the corresponding reference.

We compared HealthFound with five general-purpose language models—GPT-5.4, Gemini-3-Flash, DeepSeek-V3.2, Kimi-k2.5 and MiniMax-M2.7—as well as the Qwen3-32B backbone. To assess performance against domain-specialized models, we additionally evaluated Llama3-Med42 (70B)^34^, HuatuoGPT2 (13B and 34B)^35^, MedGemma (4B-IT and 27B-Text-IT)^36^, HuatuoGPT-o1-8B^32^ and Baichuan-M2-32B^37^ (**Supplementary Table 1**).

For disease prediction, we also compared HealthFound with Delphi-2M^20^, XGBoost^38^ and logistic regression. For numerical measurement inference, conventional baselines included mean imputation and multiple imputation by chained equations (MICE)^39^.

### UK Biobank incident disease prediction

We constructed 1,074 incident-disease endpoints by adapting FinnGen Release 12 definitions^16^ to UK Biobank. Disease ascertainment primarily used inpatient ICD-10 diagnoses and their first-recorded dates (fields 41270 and 41280), with death-cause records (fields 40001 and 40002) additionally used for mortality-related endpoints. We applied FinnGen inclusion, exclusion and composite rules, together with relevant age- and sex-specific criteria. ICD-10 subcodes were matched by prefix, and codes exceeding UK Biobank coding resolution were shortened to four characters. After manual review, duplicate or unsupported endpoint definitions were removed. All endpoints requiring unavailable surgical, medication, or reimbursement information, or unsupported disease characteristics, were excluded unless these features could be identified through diagnostic codes or implemented rules. Additional details on endpoint construction are provided in **Supplementary Methods**.

For each endpoint, participants with the target disease at or before baseline were excluded. Cases were first diagnosed during follow-up, whereas controls had no recorded target diagnosis; controls who died during follow-up were also excluded. Case inputs were restricted to observations before the first diagnosis, with all later records removed^40^. For each disease endpoint, we targeted 5,000 cases and 5,000 controls when at least 5,000 cases were available. Otherwise, all available cases were retained before feature filtering, with up to 200 additional controls selected relative to the case count, subject to availability. Within the training partition, cases and controls were balanced, with a maximum of 1,000 unique participants per class per endpoint. The training and test sets were disjoint, with all examples from each participant confined to a single partition.

The test set comprised 203,777 examples across 1,074 endpoints. We calculated accuracy, positive-class F1, sensitivity and specificity for each endpoint and summarized performance by endpoint and clinical chapter. We also evaluated low-prevalence settings using observed case proportions of 0.05%–5%. To assess how HealthFound’s positive-case detection rate varies with the time interval before disease onset across disease chapters, we conducted a stratified analysis of UK Biobank disease prediction results. For each positive case, the latest date extracted from the input text was used as the prediction start date, and the interval to the target disease onset was calculated. We then divided intervals from 0 to 14 years into seven consecutive 2-year bins, calculated the positive-case detection rate for each disease within each bin, and averaged these rates equally across diseases within each chapter. The resulting chapter-level curves were plotted against the upper boundary of each interval.

### Longitudinal health-event generation

We used HealthFound to generate possible future health-event trajectories from a participant’s longitudinal health history. Rather than adding a separate trajectory-prediction head, we formulated this as a text-generation task: the model received a participant’s historical health profile and generated a sequence of predicted future events and their occurrence times. We compared HealthFound with Delphi-2M^20^ and other LLMs using 10,000 UK Biobank participants.

#### Construct the input history

We sampled 10,000 unique participants and arranged their recorded diagnoses, assessment visits and measurements chronologically. Each participant’s history was divided at a prediction cut-off, defined as the upper median observation date and no later than the second-to-last date. For HealthFound, the historical observations were represented as a natural-language health profile.

#### Generate future trajectories

Given the historical profile, HealthFound generated possible sequences of future disease events and their predicted occurrence times. Each trajectory represented one possible continuation of the participant’s health history. We evaluated eight valid trajectories per participant for all models. For comparison, predicted events were represented using three-character ICD-10 categories and a separate death event.

#### Compare predictions with recorded events

We constructed the reference trajectory from the first recorded occurrences of ICD-10 categories absent from the input history, together with death. We then matched predicted and recorded events by their codes. Recall measured how many distinct recorded events were recovered by at least one of the eight trajectories. Because follow-up was incomplete for some participants, unrecorded events were not treated as confirmed negatives.

### Inference of missing measurements

We evaluated whether HealthFound could predict a missing biomedical measurement from the remaining information in a participant’s health profile. The evaluation covered different indicator domains, including haematology, anthropometry, biochemistry and polygenic risk scores (PRSs). The test set contained approximately 1,900 held-out examples.

For each example, we first selected one recorded measurement and removed its value from the health profile. The original value was retained only as the reference answer, while the remaining observations provided the context for prediction. We then provided the incomplete health profile to the model and asked it to infer the missing measurement. The task instruction specified the target variable and the required answer format, which could be a numerical value, categorical label or value interval, depending on the variable type.

All models used the same answer-parsing and unit-conversion rules. For numerical measurements, we compared each prediction with its recorded value using relative error. A prediction was considered correct if its relative error fell within a specified tolerance of 0.1%, 0.5%, 1%, 2%, 3%, 4% or 5%. **Supplementary Table 3** reports the comparisons at tolerances from 1% to 5%. Categorical predictions were evaluated by exact matching after code normalization, whereas interval predictions were scored by the overlap between predicted and reference ranges. These scores were evaluated separately from numerical accuracy. The scoring rules are defined in **Supplementary Methods**.

We additionally compared numerical predictions with conventional mean imputation and multiple imputation by chained equations (MICE)^39^. Results by indicator domain are reported in **Supplementary Table 12**.

### Public medical benchmarks

To assess whether HealthFound retained general medical knowledge and reasoning abilities after cohort-specific training, we evaluated it on 12,333 questions from eight public benchmarks: PubMedQA^41^, MedQA, MedXpertQA^42^, MedExQA^43^, Medbullets-opt4, Medbullets-opt5^44^, MedMCQA^45^ and MMLU-Med^46^. These benchmarks covered medical knowledge, clinical reasoning, medical examinations and literature comprehension. All models received the same questions using task-specific prompts. We extracted each model’s final answer and compared it with the reference answer to calculate accuracy for each benchmark. Overall performance was calculated as the equally weighted mean of the eight benchmark accuracies, ensuring that benchmarks with more questions did not contribute disproportionately to the final score. Benchmark descriptions and question counts are given in **Supplementary Table 7**. Data access and licensing information is given in **Supplementary Table 8**. The aggregation of benchmark scores is described in **Supplementary Methods**.

### Training-stage comparisons

To assess the contribution of each training stage, we compared the original Qwen3-32B model with HealthFound after curriculum SFT, RFT and HFPO. The three stages used approximately 10.49 million, 0.089 million, and 1.86 million training examples, respectively. These models were evaluated on the same test sets using identical prompts, decoding settings and answer-parsing rules. We also compared alternative training strategies. Independently trained standard UK Biobank SFT only used UKB disease prediction data from three stages; the replay-based training further fine-tuned the SFT model on approximately 0.089 million replayed SFT examples; and a conventional GRPO applied after SFT and RFT, with its training-data matched to that of HFPO. To distinguish the effect of staged training from that of data composition, we additionally trained single-stage SFT models, one on mixed data from SFT and RFT, and the other on mixed SFT, RFT, and HFPO data. These comparisons allowed us to examine whether performance improvements were associated with the training strategy rather than simply the amount of data used.

For measurement inference, the training-stage comparison included numerical, categorical and interval predictions, whereas the main numerical-inference evaluation considered numerical values only. Detailed results are provided in **Supplementary Table 4**.

### Literature-derived question answering

We evaluated the model’s ability to answer scientific questions using 3,574 held-out question–answer pairs derived from biomedical publications. The prompt used for data construction is provided in **Supplementary Table 13**. All models received the same questions and prompts, and their responses were compared with the corresponding reference answers.

We first measured agreement with the reference wording using BLEU-4, ROUGE-2 F1, ROUGE-L F1^47^, and word-level precision, recall and F1. Each metric was calculated for individual questions. We additionally used three independent LLM judges—GPT-5.4, DeepSeek-V4-Pro and Gemini-3-Pro-Preview—to evaluate answer correctness. The judges assessed whether each response accurately described the scientific relationship, its direction and the supporting reasoning. Their scores were averaged to obtain the final answer-correctness score.

Detailed definitions of the text-overlap metrics are provided in **Supplementary Methods**, and judge-specific results are reported in **Supplementary Table 5**.

### Disease representations and feature masking

We investigated whether cohort-specific training changed how the model represented diseases and which clinical information it used for prediction.

We compared disease embeddings extracted from HealthFound and the original Qwen3-32B model, with both models kept fixed. Each disease was represented by its endpoint name followed by the text “UK Biobank disease endpoint”. We used mean-pooled input-token embeddings from the input embedding layer to tokenize this text and averaged the corresponding token embeddings to obtain a 5,120-dimensional vector for each disease. The vectors were centred, projected to 96 dimensions, and then reduced to two dimensions using principal component analysis (PCA). The resulting plots were coloured by clinical chapter to examine whether related diseases formed coherent groups.

We further examined the predictive contribution of these features by removing selected biomedical variables from the input profiles and comparing disease-prediction accuracy with that obtained using complete profiles. These analyses characterized the model’s learned representations and use of clinical information^48^. **Supplementary Methods** describes the representation and masking analyses.

### Temporal perturbation and individual examples

To assess whether HealthFound was sensitive to temporal changes, we modified clinical event dates while preserving the original chronological order of events and keeping the measurements unchanged. We then repeated disease prediction using the modified profiles and compared accuracy with that obtained from the original records. This analysis examined whether prediction performance depended on the timing of health observations. We also examined three individual test-set examples to illustrate how HealthFound used longitudinal information for disease prediction, clinical reasoning and treatment selection. These examples were presented for qualitative interpretation. Further details are provided in **Supplementary Methods**.

### External evaluation in MIMIC-IV and NHANES

To evaluate whether HealthFound generalized beyond UK Biobank, we tested the trained model on two independent datasets. MIMIC-IV provided longitudinal hospital records for predicting future diagnoses, whereas NHANES provided cross-sectional health measurements for identifying diseases already present at the survey.

We used MIMIC-IV v3.1^26^, comprising hospitalization records from 50,880 individuals with a median observation span of 1.5 years. We constructed 408 incident-disease prediction tasks using information available by the end of each participant’s index admission. Participants with the target disease already recorded by that time were excluded. Cases had a subsequent diagnosis during follow-up, whereas controls had no recorded target diagnosis. Each task included up to 200 subjects. Model inputs included demographics, vital signs, physical measurements, laboratory results and microbiology, while target-diagnosis information and observations after the index admission were excluded. The database was collected under approvals from the institutional review boards of Beth Israel Deaconess Medical Center and Massachusetts Institute of Technology, with informed consent waived for the de-identified records.

We used health measurements from 1,534 participants in the 1999–2000 NHANES survey^27^ to construct nine prevalent-disease classification tasks. Each task included up to 200 subjects. Inputs comprised demographic, examination, questionnaire and laboratory information, excluding self-reported variables used to define each target disease. In a separate sensitivity analysis, we also removed medication- and treatment-related variables associated with the target diseases. NHANES was approved by the National Center for Health Statistics Research Ethics Review Board, and participants provided written informed consent. **Supplementary Methods** describes the external evaluation. Accuracies after removing medication and treatment variables are in **Supplementary Table 14**.

### Statistical analysis

To assess the statistical significance of HealthFound’s performance relative to baseline models, we analyzed prediction records across 1,074 UK Biobank disease endpoints. Overall accuracy was calculated by first determining accuracy for each disease and then averaging equally across all endpoints. We employed a paired cluster bootstrap using 78,316 unique participant EIDs as resampling units, whereby each bootstrap replicate sampled participants with replacement while retaining all disease-specific records belonging to each selected participant, thereby preserving within-participant correlations. For each replicate, disease-specific accuracies and their equally weighted means were recalculated to estimate the accuracy differences between HealthFound and each baseline model. A total of 5,000,000 bootstrap replicates were performed, with 95% confidence intervals derived from the 2.5th and 97.5th percentiles of the bootstrap distributions and two-sided empirical p-values calculated from the null-centered distributions. HealthFound achieved significantly higher accuracy than all baseline models, with empirical p-values reaching the Monte Carlo resolution limit of approximately 2.0 × 10⁻⁷ in every comparison.

## References

1 Saab, K. et al. Advancing conversational diagnostic AI with multimodal reasoning. Nat Med 32, 1726–1736 (2026). 10.1038/s41591-026-04371-0

2 Tu, T. et al. Towards Generalist Biomedical AI. NEJM AI 1, AIoa2300138 (2024). 10.1056/AIoa2300138

3 Singhal, K. et al. Large language models encode clinical knowledge. Nature 620, 172–180 (2023). 10.1038/s41586-023-06291-2

4 Singhal, K. et al. Toward expert-level medical question answering with large language models. Nat Med 31, 943–950 (2025). 10.1038/s41591-024-03423-7

5 Luo, R. et al. BioGPT: generative pre-trained transformer for biomedical text generation and mining. Brief Bioinform 23, bbac409 (2022). 10.1093/bib/bbac409

6 Van Veen, D. et al. Adapted large language models can outperform medical experts in clinical text summarization. Nat Med 30, 1134–1142 (2024). 10.1038/s41591-024-02855-5

7 Qiu, P. et al. Towards building multilingual language model for medicine. Nat Commun 15, 8384 (2024). 10.1038/s41467-024-52417-z

8 Zhang, K. et al. A generalist vision-language foundation model for diverse biomedical tasks. Nat Med 30, 3129–3141 (2024). 10.1038/s41591-024-03185-2

9 Savage, T., Nayak, A., Gallo, R., Rangan, E. & Chen, J. H. Diagnostic reasoning prompts reveal the potential for large language model interpretability in medicine. NPJ Digit Med 7, 20 (2024). 10.1038/s41746-024-01010-1

10 Tu, T. et al. Towards conversational diagnostic artificial intelligence. Nature 642, 442–450 (2025). 10.1038/s41586-025-08866-7

11 Wornow, M. et al. The shaky foundations of large language models and foundation models for electronic health records. NPJ Digit Med 6, 135 (2023). 10.1038/s41746-023-00879-8

12 Jensen, A. B. et al. Temporal disease trajectories condensed from population-wide registry data covering 6.2 million patients. Nat Commun 5, 4022 (2014). 10.1038/ncomms5022

13 Price, N. D. et al. A wellness study of 108 individuals using personal, dense, dynamic data clouds. Nat Biotechnol 35, 747–756 (2017). 10.1038/nbt.3870

14 Bycroft, C. et al. The UK Biobank resource with deep phenotyping and genomic data. Nature 562, 203–209 (2018). 10.1038/s41586-018-0579-z

15 Sudlow, C. et al. UK biobank: an open access resource for identifying the causes of a wide range of complex diseases of middle and old age. PLoS Med 12, e1001779 (2015). 10.1371/journal.pmed.1001779

16 Kurki, M. I. et al. FinnGen provides genetic insights from a well-phenotyped isolated population. Nature 613, 508–518 (2023). 10.1038/s41586-022-05473-8

17 Rajkomar, A. et al. Scalable and accurate deep learning with electronic health records. NPJ Digit Med 1, 18 (2018). 10.1038/s41746-018-0029-1

18 Guo, L. L. et al. A multi-center study on the adaptability of a shared foundation model for electronic health records. NPJ Digit Med 7, 171 (2024). 10.1038/s41746-024-01166-w

19 Kraljevic, Z. et al. Foresight-a generative pretrained transformer for modelling of patient timelines using electronic health records: a retrospective modelling study. Lancet Digit Health 6, e281–e290 (2024). 10.1016/S2589-7500(24)00025-6

20 Shmatko, A. et al. Learning the natural history of human disease with generative transformers. Nature 647, 248–256 (2025). 10.1038/s41586-025-09529-3

21 Kirchler, M. et al. Large language models improve transferability of electronic health record-based predictions across countries and coding systems. NPJ Digit Med 9, 177 (2026). 10.1038/s41746-026-02363-5

22 Hegselmann, S. et al. Large language models are powerful electronic health record encoders. NPJ Digit Med 9, 530 (2026). 10.1038/s41746-026-02915-9

23 Sivarajkumar, S. et al. A multimodal generative model for structured and unstructured electronic health records. Npj Health Syst 3, 46 (2026). 10.1038/s44401-026-00095-y

24 Kirkpatrick, J. et al. Overcoming catastrophic forgetting in neural networks. Proc Natl Acad Sci U S A 114, 3521–3526 (2017). 10.1073/pnas.1611835114

25 Guo, D. et al. DeepSeek-R1 incentivizes reasoning in LLMs through reinforcement learning. Nature 645, 633–638 (2025). 10.1038/s41586-025-09422-z

26 Johnson, A. E. W. et al. MIMIC-IV, a freely accessible electronic health record dataset. Sci Data 10, 1 (2023). 10.1038/s41597-022-01899-x

27 Zipf, G. et al. National health and nutrition examination survey: plan and operations, 1999-2010. Vital Health Stat 1 (56), 1–37 (2013).

28. Wang, Z. et al. A world model simulates the latent dynamics of human health. Preprint at https://www.medrxiv.org/content/10.64898/2026.09.19.26363460v2 (2026).

29 Fry, A. et al. Comparison of Sociodemographic and Health-Related Characteristics of UK Biobank Participants With Those of the General Population. Am J Epidemiol 186, 1026–1034 (2017). 10.1093/aje/kwx246

30 Vasey, B., et al. Reporting guideline for the early stage clinical evaluation of decision support systems driven by artificial intelligence: DECIDE-AI. BMJ 377, e070904 (2022). 10.1136/bmj-2022-070904

31 Kwon, W. et al. Efficient Memory Management for Large Language Model Serving with PagedAttention. in Proceedings of the 29th Symposium on Operating Systems Principles 611–626 (ACM, 2023).

32. Chen, J. et al. Towards Medical Complex Reasoning with LLMs through Medical Verifiable Problems. in Findings of the Association for Computational Linguistics: ACL 2025 14552–14573 (Association for Computational Linguistics, 2025).

33 Lambert, N. et al. Tülu 3: Pushing Frontiers in Open Language Model Post-Training. in Conference on Language Modeling Vol. 2 (COLM, 2025).

34. Christophe, C., Kanithi, P. K., Raha, T., Khan, S. & Pimentel, M. A. F. Med42-v2: A Suite of Clinical LLMs. Preprint at https://arxiv.org/abs/2408.06142 (2024).

35. Chen, J. et al. HuatuoGPT-II, One-stage Training for Medical Adaption of LLMs. in Conference on Language Modeling Vol. 1 (COLM, 2024).

36 Sellergren, A. et al. An open vision-language model for diverse medical applications. Nat Med 32, 3695–3704 (2026). 10.1038/s41591-026-04626-w

37. Dou, C., et al. Baichuan-M2: Scaling Medical Capability with Large Verifier System. Preprint at https://arxiv.org/abs/2509.02208 (2025).

38 Chen, T. & Guestrin, C. XGBoost: A Scalable Tree Boosting System. in Proceedings of the 22nd ACM SIGKDD International Conference on Knowledge Discovery and Data Mining 785–794 (ACM, 2016).

39 White, I. R., Royston, P. & Wood, A. M. Multiple imputation using chained equations: Issues and guidance for practice. Stat Med 30, 377–399 (2011). 10.1002/sim.4067

40 Kapoor, S. & Narayanan, A. Leakage and the reproducibility crisis in machine-learning-based science. Patterns (N Y) 4, 100804 (2023). 10.1016/j.patter.2023.100804

41 Jin, Q., Dhingra, B., Liu, Z., Cohen, W. & Lu, X. PubMedQA: A Dataset for Biomedical Research Question Answering. in Proceedings of the 2019 Conference on Empirical Methods in Natural Language Processing and the 9th International Joint Conference on Natural Language Processing (EMNLP-IJCNLP) 2567–2577 (Association for Computational Linguistics, 2019).

42 Zuo, Y., et al. MedXpertQA: Benchmarking Expert-Level Medical Reasoning and Understanding. in Proceedings of the 42nd International Conference on Machine Learning Vol. 267 80961–80990 (PMLR, 2025).

43 Kim, Y., Wu, J., Abdulle, Y. & Wu, H. MedExQA: Medical Question Answering Benchmark with Multiple Explanations. in Proceedings of the 23rd Workshop on Biomedical Natural Language Processing 167–181 (Association for Computational Linguistics, 2024).

44 Chen, H., Fang, Z., Singla, Y. & Dredze, M. Benchmarking Large Language Models on Answering and Explaining Challenging Medical Questions. in Proceedings of the 2025 Conference of the Nations of the Americas Chapter of the Association for Computational Linguistics: Human Language Technologies (Volume 1: Long Papers) 3563–3599 (Association for Computational Linguistics, 2025).

45 Pal, A., Umapathi, L. K. & Sankarasubbu, M. MedMCQA: A Large-scale Multi-Subject Multi-Choice Dataset for Medical domain Question Answering. in Proceedings of the Conference on Health, Inference, and Learning Vol. 174 248–260 (PMLR, 2022).

46. Hendrycks, D. et al. Measuring Massive Multitask Language Understanding. in International Conference on Learning Representations (ICLR, 2021).

47 Zhao, M. et al. Automating evaluation of LLM-generated responses to patient questions about rare diseases. JAMIA Open 9, ooag054 (2026). 10.1093/jamiaopen/ooag054

48 Castro, D. C., Walker, I. & Glocker, B. Causality matters in medical imaging. Nat Commun 11, 3673 (2020). 10.1038/s41467-020-17478-w

